# Taxane-based Chemotherapy is Effective in Metastatic Appendiceal Adenocarcinoma

**DOI:** 10.1101/2023.07.11.23292522

**Authors:** Julia Dansby, Aditya More, Mohammad Zeineddine, Abdelrahman Yousef, Alisha Bent, Farshid Dayyani, Robert Wolff, Michael Overman, John Paul Shen

## Abstract

Appendiceal cancer is a rare, orphan disease with no therapies currently approved by the FDA for its treatment. Given the limited data regarding drug efficacy, these tumors have historically been treated with chemotherapy designed for colon cancer. However, an overwhelming body of molecular data has demonstrated that appendiceal adenocarcinoma is a distinct entity with key molecular differences from colon cancer, notably rare *APC* mutation. Recognizing that APC loss-of-function is thought to contribute to taxane resistance, and that taxanes are effective in the treatment of other gastrointestinal tumors including gastric, esophageal, and small bowel adenocarcinoma, we completed a single-center retrospective study to assess efficacy. In a cohort of 13 patients with metastatic appendiceal adenocarcinoma, treated with taxane chemotherapy the median overall survival was 8.3 months. Of 10 evaluable patients we observed 3 responses, 4 patients with stable disease, and 3 with progression (30% response rate, 70% disease control rate). The results of this study showing activity of taxane-based chemotherapy in appendiceal adenocarcinoma support further clinical investigation of taxane therapy in this orphan disease.

## Introduction

Appendiceal tumors encompass a rare and diverse group of neoplasms, with appendiceal adenocarcinoma (AA) being the most common histologic subtype^1^. Despite a unique natural history characterized by metastatic spread limited to the peritoneum, as well as growing evidence that appendiceal tumors are molecularly distinct from colorectal cancer (CRC)^2^, current National Comprehensive Cancer Network (NCCN) guidelines suggest that appendiceal tumors should be treated with the same chemotherapy as used for CRC. However, recent prospective and retrospective studies have called into question the dogma that appendiceal tumors respond to chemotherapy similarly to colon cancer^3,4^. In particular a prospective, randomized, crossover design trial showed that patients with low-grade mucinous appendiceal adenocarcinomas do not derive benefit from 5-FU-based chemotherapy^4^, highlighting the need to drug development efforts specific to appendix cancer.

Taxanes have been shown to be ineffective in colorectal cancer but are active in small bowel adenocarcinoma^5^. It has been reported that *APC* loss-of-function is a mechanism of taxane resistance^6^. Unlike CRC, where *APC* is mutated in >70% of tumors, *APC* mutation is uncommon in subtypes of appendiceal cancer (9.0%)^2^. We therefore hypothesized that taxane chemotherapy would have activity in appendiceal adenocarcinoma.

## Materials and Methods

The MD Anderson adapted version of the Palantir-Foundry software system was used to perform an automated query of the MD Anderson GI Medical Oncology database to identify patients with appendiceal adenocarcinoma treated with paclitaxel-based regimens between 2003 – 2022. This activity was approved by the University of Texas MD Anderson Cancer Center Institutional Review Board (IRB) under protocol Lab09-0373. Manual chart review was performed to confirm patients met eligibility criteria and to extract outcome variables. Eligible patients had pathologic diagnosis of appendiceal adenocarcinoma, mucinous adenocarcinoma, signet ring cell adenocarcinoma, or goblet cell adenocarcinoma, and more than one dose of taxane-based therapy and were not enrolled in a clinical trial. Radiographic response was assessed retrospectively based upon the treating physician’s assessment and categorized as response, stable disease, progression; response could not be evaluated (NA) if the patient did not have imaging performed after starting taxane therapy. Biochemical response was evaluated based on the percent change in tumor marker before and after treatment. Patients with a greater than 20% decrease were categorized as response, less than 20% change as stable, and greater than 20% increase as progression. Those whose respective tumor markers were not elevated were indicated as such, and those whose tumor markers were not measured were classified as NA. Median overall survival (OS) was determined using the Kaplan-Meier method.

## Results

Thirteen patients with appendiceal adenocarcinoma treated with paclitaxel-based therapy were identified and met inclusion criteria. Median age was 64 years (range 29-77) with roughly equal splits between male and female as well as well, moderate, and poorly differentiated tumors (**Supplemental Table 1**). All 13 patients had peritoneal disease at time of treatment, 7 (54%) had prior surgical resection. The cohort was heavily pretreated with median of 3 prior lines of therapy (range 1-6), 10 of the 13 had prior treatment with either FOLFOX of CapeOx. Ten patients received taxane-based combinations (gemcitabine combination in 7, platinum combination in 2, and fluoropyrimidine combination in 1), 3 received paclitaxel monotherapy. Four of the combination treated patients received nab-paclitaxel, the rest paclitaxel.

The median OS from the start of taxane therapy was 8.77 months (range 0.7 – 31.0 months, **Figure 1A**) with 3 of 13 patients still alive at time of analysis. Four patients stopped therapy after 3 or fewer taxane treatments due to either Small Bowel Obstruction (SBO) or deteriorating performance status, median Progression Free Survival (PFS) for the remaining 9 patients was 7.4 months (range 0.8 – 15.5 months, **Figure 1B)**. Median time on treatment was 2.5 months (range 0.2 – 13.1 months, **Figure 1C**) with three patients remaining on treatment at time of analysis. Of the 10 patients that could be assessed for radiographic response, 3 showed response, 4 with stable disease, and 3 with progression for a response rate of 30% and a disease control rate of 70% (**Figure 1C**). Of the 9 patients with elevated CEA, biochemical response was seen in 3 (33%), stable disease in 3 (33%), and progression in 3 (33%). Of the 6 patients with elevated CA 19-9, biochemical response was seen in 2 (33%), stable disease in 2 (33%), and progression in 2 (33%). Two patients received first line carboplatin and paclitaxel, both had response. One patient was initially diagnosed with a primary ovarian tumor and changed to 5-FU and bevacizumab after 3 cycles once expert pathology review made diagnosis of appendiceal adenocarcinoma. The second patient had concurrent diagnoses of stage III squamous cell carcinoma of the lung and goblet cell adenocarcinoma of the appendix and was treated initially with weekly carboplatin and paclitaxel with concurrent radiation and then carboplatin and paclitaxel plus pembrolizumab per NSCLC guidelines. Treatment with carboplatin and paclitaxel is ongoing, with marked radiographic improvement in peritoneal carcinomatosis and complete response of NSCLC based on imaging and endobronchial biopsies. There was not an associated between grade and radiographic response. Three of four GNAS mutant tumors did not respond, consistent with prior report suggesting intrinsic resistance to therapy in the case of mutant *GNAS*^3^.

**Figure 1.**
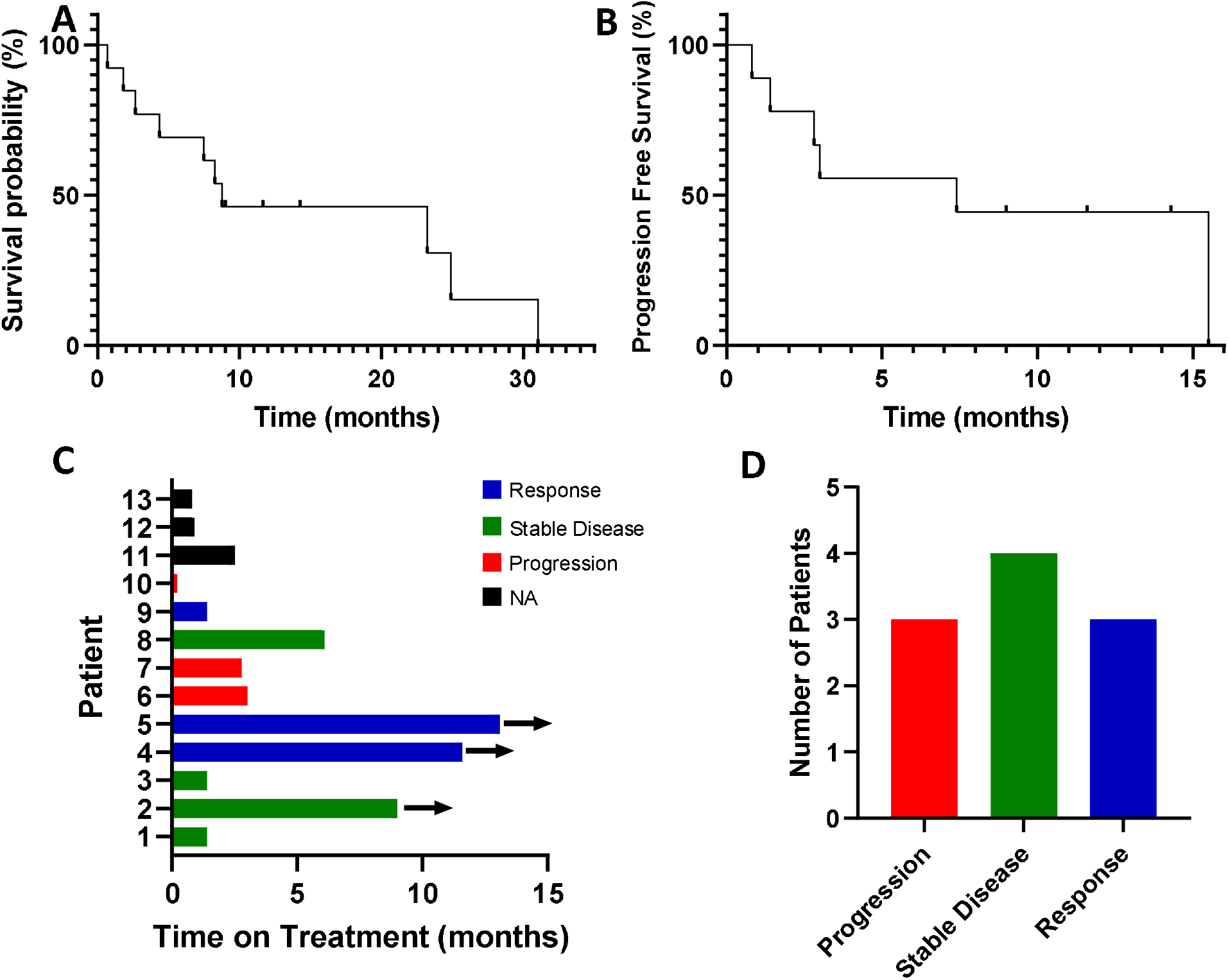
Outcomes for AA patients treated with taxane-based therapy. Overall **(A)** and Progression Free **(B)** Survival from the start of treatment. **(C)** Time on treatment, arrows indicate patient remains on treatment. **(D)** Radiographic response.

## Discussion

Complete cytoreductive surgery (CRS) with Heated Intraperitoneal Chemotherapy (HIPEC) remains the treatment of choice for patients with metastatic appendiceal adenocarcinoma^7^, however for many patients the extend of peritoneal metastatic disease precludes this treatment option. There is unfortunately very little prospective data to guide chemotherapy choice for these non-operative patients. To our knowledge, this is the largest cohort of appendiceal cancer patients treated with taxane-based therapy reported in the literature. Although we recognize the inherent limitations of this small, retrospective study design, the favorable disease control rate in such a heavily pretreated cohort indicates that taxane-based chemotherapy is active in appendiceal adenocarcinoma and should be further studied in a prospective fashion. These findings are consistent with the activity of taxanes in small bowel adenocarcinoma (SBA)^5 8^, are also consistent with the activity of intraperitoneal (IP) injection of paclitaxel in orthotopic PDX models of appendiceal cancer^9^ and activity of IP paclitaxel in gastric and ovarian cancers^10^.

**Table 1.**

| Patient | Line of Therapy | Regimen | CEA Response | CA 19-9 Response | Radiographic Response | Histology | Grade | Stage | Signet Ring Features | CRS/ HIPEC | Mutation Status |
| --- | --- | --- | --- | --- | --- | --- | --- | --- | --- | --- | --- |
| 1 | 3 | Gemcitabine + paclitaxel | Not Elevated | N/A | SD | Mucinous Adenocarcinoma | Poorly Differentiated | IV | Y | N | None |
| 2 | 6 | Paclitaxel | SD | SD | SD | Mucinous Adenocarcinoma | Well Differentiated | IV | N | N | None |
| 3 | 2 | Gemcitabine + paclitaxel | SD | SD | SD | Mucinous Adenocarcinoma | Moderately Differentiated | IV | N | N | U2AF1, KRAS |
| 4 | 3 | Gemcitabine + nab-paclitaxel | Response | Response | Response | Adenocarcinoma | Moderately Differentiated | IV | N | Y | KRAS, GNAS, CHEK2, TP53 |
| 5 | 1 | Carboplatin + pembrolizumab + paclitaxel | Not Elevated | Not Elevated | Response | Goblet Cell Adenocarcinoma | Poorly Differentiated | IV | N | N | CTNNB1, NF1, TP53, FBXW7, RET |
| 6 | 3 | Gemcitabine + paclitaxel | Response | Not Elevated | Progression | Adenocarcinoma | Moderately Differentiated | IV | N | N | GNAS |
| 7 | 3 | Paclitaxel | SD | Response | Progression | Mucinous Adenocarcinoma | Poorly Differentiated | IV | Y | N | TP53, KRAS |
| 8 | 5 | Gemcitabine + capecitabine + nab-Paclitaxel | Response | N/A | SD | Mucinous Adenocarcinoma | Moderately Differentiated | IV | N | Y | CTNNB1, KRAS |
| 9 | 1 | Carboplatin + paclitaxel | N/A | N/A | Response | Mucinous Adenocarcinoma | Poorly Differentiated | IV | Y | N | None |
| 10 | 3 | Paclitaxel | Progression | Not elevated | Progression | Mucinous Adenocarcinoma | Poorly Differentiated | IV | Y | N | GNAS |
| 11 | 3 | Gemcitabine + paclitaxel | Progression | Progression | N/A | Mucinous Adenocarcinoma | Well Differentiated | IV | N | Y | CREBBP, KRAS, GNAS, SMARCA4 |
| 12 | 5 | Gemcitabine + nab-paclitaxel | N/A | N/A | N/A | Mucinous Adenocarcinoma | Well Differentiated | IV | N | N | None |
| 13 | 4 | Fluoropyrimidine + nab-Paclitaxel | Progression | Progression | N/A | Adenocarcinoma | Moderately Differentiated | IV | N | Y | KRAS |

## Data Availability

All data other than individual patient level data are contained in the manuscript, patient level data produced in the present study are available upon reasonable request to the authors

## Author Contributions

Conception/Design- MO, JPS

Provision of study material or patients- AB, MO, FD, RW, JPS

Collection and/or assembly of data- JD, AM, MZ, AY, JPS

Data analysis and interpretation – JD, JPS

Manuscript writing – JD, JPS

Final approval of manuscript – all authors

## Conflict of Interest / Commercial Interest

Consulting/advisory relationship- JPS (Engine Biosciences, NaDeNo Nanoscience) Research Funding – JPS (NIH, ASCO, CPRIT)

Employment

Expert Testimony

Honoraria

Ownership Interests

Intellectual Property Rights/inventor/patent holder

Scientific Advisory Board- JPS (ACPMP), MO (ACPMP)

## Acknowledgements

This work was supported by the Col. Daniel Connelly Memorial Fund, the National Cancer Institute (K22 CA234406 to J.P.S., and the Cancer Center Support Grant (P30 CA016672), the Cancer Prevention & Research Institute of Texas (RR180035 to J.P.S., J.P.S. is a CPRIT Scholar in Cancer Research), and a Conquer Cancer Career Development Award to J.P.S. Any opinions, findings, and conclusions expressed in this material are those of the author(s) and do not necessarily reflect those of the American Society of Clinical Oncology® or Conquer Cancer.

---

**Supplemental Table 1:** Patients’ baseline characteristics

| Characteristic | n (%) |
| --- | --- |
| Median age at diagnosis (range), yr | 64 (29-77) |
| Female | 7 (54) |
| Male | 6 (46) |
| White | 11 (85) |
| Grade |  |
| Poor | 5 (38) |
| Moderate | 5 (38) |
| Well | 3 (23) |
| Peritoneal Disease | 13 (100) |
| Primary tumor resected | 7 (54) |
| Previous treatment with FOLFOX or XELOX | 10 |
| Previous treatment with anti-VEGF therapy | 4 |
| Microsatellite status |  |
| Microsatellite Stable | 6 (46) |
| Microsatellite Unknown | 7 (54) |
| Treatment |  |
| Combination | 10 (77) |
| Gemcitabine + nab-paclitaxel | 2 |
| Gemcitabine + paclitaxel | 4 |
| Carboplatin + paclitaxel + pembrolizumab | 1 |
| Carboplatin + paclitaxel | 1 |
| Gemcitabine + nab-paclitaxel + capecitabine | 1 |
| Fluoropyrimidine + nab-paclitaxel | 1 |
| Monotherapy | 3 (23) |
| Paclitaxel | 3 |

